# Plasma levels of soluble ACE2 are associated with sex, Metabolic Syndrome, and its biomarkers in a large cohort, pointing to a possible mechanism for increased severity in COVID-19

**DOI:** 10.1101/2020.06.10.20127969

**Authors:** Sergey A. Kornilov, Isabelle Lucas, Kathleen Jade, Chengzhen L. Dai, Jennifer C. Lovejoy, Andrew T. Magis

**Affiliations:** Institute for Systems Biology, Seattle, WA, 401 Terry Ave N, Seattle WA 98109-5263

**Keywords:** ACE2, SARS-CoV-2, COVID-19, Metabolic Syndrome, GGT, liver function

## Abstract

We examined the associations between plasma concentrations of soluble ACE2 and biomarkers of Metabolic Syndrome in a large (N=2,051) sample of individuals who participated in a commercial wellness program and who underwent deep molecular phenotyping. sACE2 levels were significantly higher in men, compared to women, and in individuals with Metabolic Syndrome, compared to controls. sACE2 levels showed reliable associations with all individuals components of Metabolic Syndrome, including obesity, hypertension, insulin resistance, hyperlipidemia, and as well as markers of liver damage. This profile of associations was statistically significantly stronger in men, compared to women, and points to preexisting cardiometabolic conditions as possible risk factors for increased severity of symptoms in some COVID-19 patients through increased expression of ACE2 in the liver.

---

Patients who are at high risk for mortality from COVID-19, the disease caused by severe acute respiratory syndrome coronavirus 2 (SARS-CoV-2), are more likely to be older and male, and to have chronic diseases such as hypertension, diabetes, cardiovascular, and chronic lung disease [1,2]. Although the mechanisms behind these associations are poorly understood, this increased risk could be partly associated with increased expression of the cellular receptor of SARS-CoV-2, angiotensin-converting enzyme-2 (ACE2), which is found at elevated levels in older individuals, men, and in cardiovascular and inflammatory conditions [3,4]. ACE2 maintains homeostasis of the renin-angiotensin system and converts angiotensin II to angiotensin 1-7, which has vasodilatory and anti-inflammatory properties. It occurs in a membrane bound form (mACE2), highly expressed in heart, airways, kidney, and liver tissue, and in an enzymatically active soluble form (sACE2) that is generated in response to inflammatory signals and disease via mACE2 shedding into the blood where it normally is found at low levels in healthy individuals.

We interrogated the associations between plasma concentrations of sACE2 and biomarkers of Metabolic Syndrome (Body Mass Index, BMI; blood pressure; glycemic markers, and lipid levels), adiposity (plasma leptin and serum adiponectin), inflammation (high-sensitivity C-reactive protein, hsCRP, white cell count, and interleukin-8) and liver damage (alanine aminotransferase, aspartate transaminase, and gamma-glutamyl-transferase, GGT) in data from a large cohort of participants in a commercial wellness program who had undergone comprehensive multi-omic profiling (N=2,051; 1,238 women and 813 men, aged 22 to 87 years, M=47.3, SD=11.71) (see [5] for details). Clinical laboratory tests were performed in CLIA-certified laboratories by Quest Diagnostics or LabCorp. Plasma sACE2 and leptin levels were measured via proximity extension immunoassaying using Olink® Cardiovascular II proteomics panel. Analyses were performed using transformed and scaled biomarker values in a robust linear regression framework while controlling for age, sex (except sex-specific analyses), eight genetic principal components, smoking, vendor, season, and use of diabetes, cholesterol-lowering, and ACE-inhibitor medications.

Confirming results from recent studies [3,4], we found higher plasma sACE2 levels in men compared to women (P=2×10^−16^), and in older individuals (P=8.6×10^−11^), with the age association more pronounced in women (for the interaction, P_int_=0.02). We found higher levels of sACE2 in post-menopausal women, compared to pre-menopausal women (P=0.02; see Figure 1).

**Figure 1.**
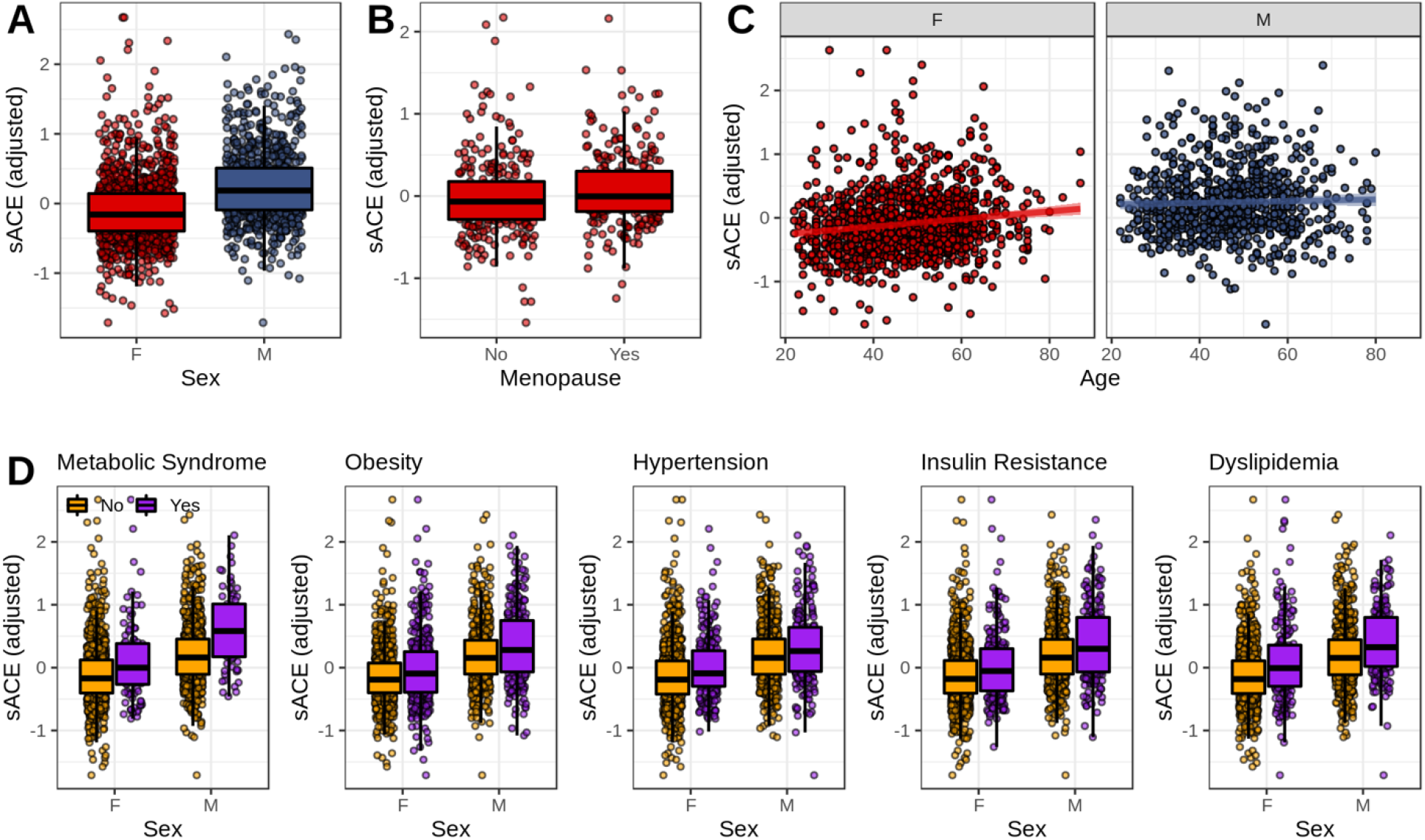
The associations of plasma sACE2 levels with sex, age, and Metabolic Syndrome. Panel A: sex differences in plasma sACE2 levels. Panel B: differences in sACE2 levels between pre- (N=272) and post-menopausal women (N=251) women over 35 years of age. Panel C: associations between sACE2 and age. Panel D: differences in sACE2 levels in individuals who do vs. do not meet diagnostic criteria for MetS and it subcomponents. The following diagnostic criteria, based on WHO guidelines, were used: Obesity: BMI>30 kg/m^2^, Hyperglycemia: fasting glucose >100 mg/dl or other evidence of insulin resistance (e.g., prescription); Hypertension: systolic/diastolic blood pressure >=140/90 mm/hg; Dyslipidemia: triglycerides > 150 or HDL-C <35 for men and <39 for women. Overall status was determined as MetS if the individual met criteria for insulin resistance and satisfied at least one other domain criterion for the syndrome. sACE2: soluble ACE2 (adjusted for covariates and scaled).

Individuals who met World Health Organization’s (WHO) diagnostic criteria for MetS (N=171) displayed elevated plasma sACE2 levels compared to controls (N=1,880; P=4.7×10^−5^), and the effect was stronger in men (P_int_=8.9×10^−5^). We found all of MetS component biomarkers to be positively associated with plasma sACE2 (see Figure 2). The associations were significantly stronger in men for biomarkers of obesity and adiposity (BMI, P_int_=0.0123; leptin, P_int_=0.0342), insulin resistance and hyperglycemia (HbA1c, P_int_=0.0368; HOMA-IR, P_int_=0.042), as well as triglycerides (P_int_=0.0134) and serum hsCRP (P_int_=0.041). Among examined biomarkers, we found the strongest association between sACE2 and GGT (P=3.44×10^−90^), an enzyme that is an important indicator of oxidative stress and liver and bile duct damage. The association between GGT and sACE2 was also preferentially stronger in men (P=0.01).

**Figure 2.**
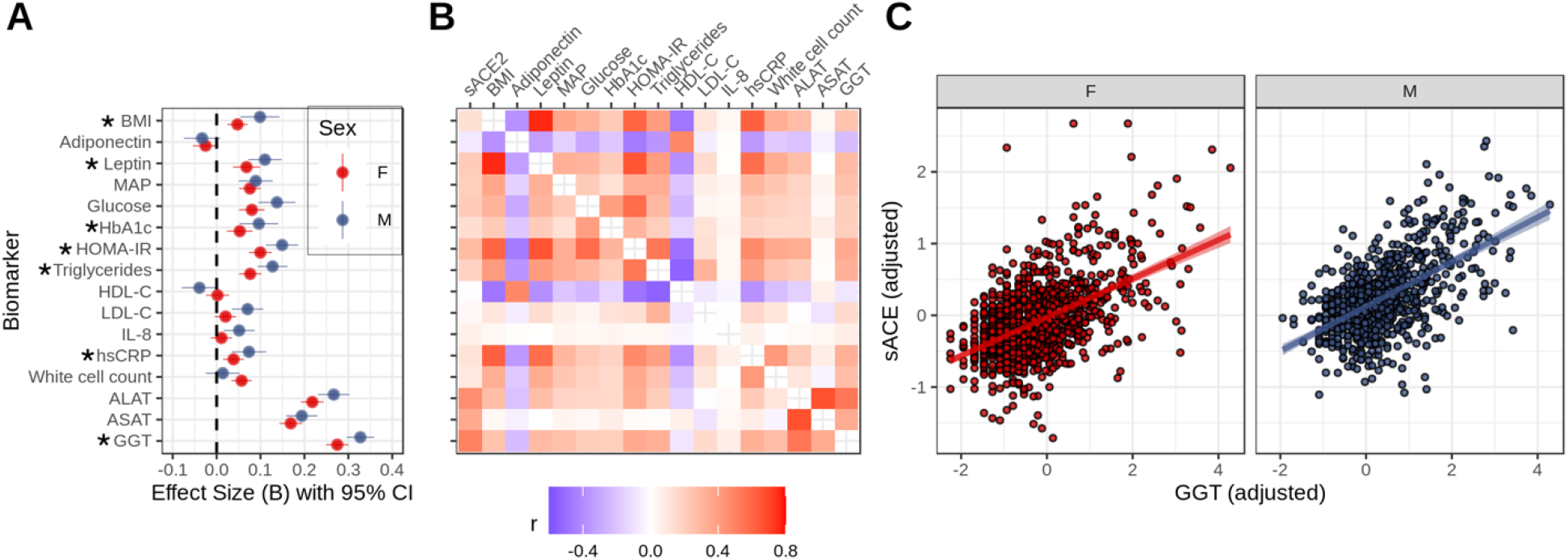
Associations between sACE2 and biomarkers of Metabolic Syndrome, inflammation, and liver damage. Panel A: effect size estimates from robust linear regressions predicting sACE2 from biomarkers estimated for men and women separately. Effect sizes are in NPX (log2 metric for the sACE2 measurement) per each standard deviation increase in transformed biomarker. Biomarkers for which a significant interaction with sex was established are marked with *. Panel B: partial Spearman correlations between study biomarkers and sACE2 levels in the total sample. Panel C: scatterplot of associations between GGT and sACE2. BMI: Body Mass Index; MAP – Mean Arterial Blood Pressure, HbA1c – glycohemoglobin A1c, HOMA-IR – homeostatic model assessment of insulin resistance, HDL-C - high-density lipoprotein cholesterol, LDL-C – low-density lipoprotein cholesterol. IL-8 - interleukin 8, hsCRP – high-sensitivity C-reactive protein, ALAT – alanine aminotransferase, ASAT – aspartate transaminase, GGT – gamma-glutamyl-transferase.

The observed robust pattern of associations between increased plasma sACE and MetS points to the possible shared pathways in cardiometabolic disease and COVID-19, implicating insulin resistance, chronic inflammation, and liver damage. This pattern is intriguing given that both sACE2 and mACE2 have been shown to be upregulated in a rat model of chronic liver disease and that sACE2 levels are higher in patients with heart failure [4]. This upregulation may be related to the tissue-specific patterns of increased SARS-CoV-2 infectivity in patients with cardiovascular disease and/or liver damage, and warrants further research on sACE2 as a potential biomarker for COVID-19 severity.

## Data Availability

The dataset supporting the conclusions of this article is available from the authors upon request.

## Declarations

### Ethics approval and consent to participate

All research was conducted in accordance to regulations and guidelines for observational research in human subjects. The study was reviewed and approved by the Western IRB (Study Number 1178906). The research was performed entirely using de-identified and aggregated data of individuals who had signed a research authorization allowing the use of their anonymized data in research. Per current U.S. regulations for use of deidentified data, informed consent was not required.

## Consent for publication

Not applicable.

## Availability of data and materials

The dataset supporting the conclusions of this article is available from the authors upon request.

## Authors’ contributions

SK, ILB, KJ, JL, CD, AM conceptualized the study and drafted the manuscript. SK, CD, AM performed data quality control and assurance, transformation, and data analysis. All authors critically revised the manuscript.

## Acknowledgements

The express their gratitude to the members of the Arivale program for granting the permission to use their deidentified data for research, and to Mr. Brett Smith at Institute for Systems Biology for his contribution to the study.

## List of abbreviations

sACE2/mACE2: soluble/membrane-bound angiotensin-converting enzyme-2
MetS: Metabolic Syndrome
GGT: gamma-glutamyl-transferase
BMI: Body Mass Index
HbA1c: glycohemoglobin A1c
HOMA-IR: homeostatic model assessment of insulin resistance
hs-CRP: high-sensitivity C-reactive protein

## References

1. Tian W, Jiang W, Yao J, et al. Predictors of mortality in hospitalized COVID-19 patients: A systematic review and meta-analysis. J Med Virol. 2020;10.1002/jmv.26050.

2. Guo W, LM, Dong Y, et al. Diabetes is a risk factor for the progression and prognosis of COVID-19. Diabetes Metab Res Rev. 2020: e3319.

3. Swärd P, Edsfeldt A, Reepalu A, Jehpsson L, Rosengren BE, Karlsson MK. Age and sex differences in soluble ACE2 may give insights for COVID-19. Crit Care. 2020;24(1):221.

4. Sama IE, Ravera A, Santema BT, et al. Circulating plasma concentrations of angiotensin-converting enzyme 2 in men and women with heart failure and effects of renin-angiotensin-aldosterone inhibitors. Eur Heart J. 2020;41(19):1810–1817.

5. Zubair N., Conomos M., Hood L., et al. Genetic Predisposition Impacts Clinical Changes in a Lifestyle Coaching Program. Sci Rep 9. 2019:6805.

6. Herath CB., Warner, FJ., et al. Upregulation of hepatic Upregulation of hepatic angiotensin-converting enzyme 2 (ACE2) and angiotensin-(1–7) levels in experimental biliary fibrosis. Journ of Hepatol. 2007;47(3):387–395

